# Association of mid-age Life’s Essential 8 score with digital cognitive performance and incident Alzheimer’s disease: the Framingham Heart Study

**DOI:** 10.1101/2024.05.06.24306950

**Authors:** Jian Yang, Huitong Ding, Yi Li, Ting Fang Alvin Ang, Sherral Devine, Yulin Liu, Wendy Qiu, Rhoda Au, Jiantao Ma, Chunyu Liu

## Abstract

**Background:** Emerging studies indicate that cardiovascular health (CVH) is a modifiable risk factor for AD. However, studies on how mid-life Life’s Essential 8 (LE8) scores affect the AD risk and digital cognitive performance are limited.

**Objective:** To examine the associations between CVH in middle age, as defined by LE8 scores, and subsequent digital cognitive performance and incident AD.

**Methods:** Linear regression and Cox proportional-hazard models were used to examine the associations of mid-age CVH with dCDT performance and incident AD, respectively. Prediction performance of mid-age LE8 scores for incident AD were assessed using ROC curve analysis.

**Results:** Every one-SD increase in mid-age LE8 total score was associated with a 0.16-SD increase in dCDT total score (p < 0.001). Ideal CVH in middle age was associated with higher dCDT scores compared to intermediate CVH. Furthermore, higher mid-age LE8 scores were associated to a decreased risk of AD, with ideal CVH significantly lowering AD risk compared to intermediate CVH. The combination of dCDT performance, mid-age LE8 scores, APOE ε4 status, and other covariates provided the best prediction performance for incident AD, with an AUC of 0.84. Notably, mid-life LE8 scores improved its predictive accuracy by 5.7%.

**Conclusion:** Our findings emphasize the critical role of CVH in middle age as a predictor of both digital cognitive performance and the risk of developing AD, highlighting the importance of early intervention on CVH to delay the progression of cognitive decline.

## INTRODUCTION

Alzheimer’s disease (AD) poses a significant challenge to public health due to its gradual onset and the progressive deterioration of cognitive functions [1]. With effective treatments for AD being scarce, the focus has shifted towards early detection as a key strategy for managing the disease [2]. Studies highlight the importance of mitigating major risk factors, particularly cardiovascular health (CVH), during middle age before cognitive decline begins, to lower AD risk [3–6]. The American Heart Association (AHA) has put forward Life’s Simple 7 (LS7) as a CVH metric targeting modifiable risk factors including blood pressure, blood sugar, cholesterol levels, body mass index (BMI), physical activity, diet, and smoking [7]. Many studies have reported a significant association between higher LS7 scores and decreased AD risk [8–10]. Building on LS7, the AHA has recently introduced Life’s Essential 8 (LE8), refining the model to incorporate sleep quality and a 0 to 100 scale for a more precise CVH quantification [11]. Despite this, the effect of mid-life LE8 scores on cognitive health and their influence on AD risk remains an underexplored area.

Understanding the association between mid-life LE8 scores and early cognitive decline requires accurate tracking of these initial subtle cognitive changes. Advances in technology have led to the development of precise digital methods for cognitive function assessment, significantly surpassing traditional neuropsychological (NP) tests in terms of objectivity and sensitivity to early signs of cognitive decline. The digital Clock Drawing Test (dCDT) exemplifies such innovation by employing a digitizing ballpoint pen and grid-patterned paper to replicate the conventional clock drawing test [12]. This approach utilizes advanced software to track pen movements, thus refining CDT and NP evaluations with standardized administration, objective scoring, and reduced biases. Additionally, dCDT covers a broad spectrum of cognitive domains and utilizes a comprehensive scoring system for an in-depth analysis of cognitive abilities [12, 13]. Therefore, determining the association between mid-life CVH, as indicated by LE8 scores, and dCDT’s capacity for early cognitive assessment is essential for timely intervention strategies.

The objective of this study was to investigate associations of mid-age LE8 scores with dCDT and incident AD in the Framingham Heart Study (FHS), aiming to offer essential insights for the prevention of cognitive decline.

## METHODS

### Study population

The FHS, established in 1948, is a community-based cohort study [14]. The Offspring Cohort of the FHS began in 1971, registering 5,124 individuals, including children of the Original Cohort and their spouses. These participants are subject to routine health examinations roughly every four years, with ten such examinations conducted so far, during which a broad array of health data is gathered [15, 16].

This study included 1,413 participants from the FHS Offspring Cohort, each of whom had participated in at least one dCDT since 2011 and had been involved in at least one of the FHS core exams from 5 to 9 (1991-2014). Exclusions were made for participants under 60 years of age (n=67) and those diagnosed with non-AD dementia (n=25) at the time of dCDT. An additional 123 participants were excluded from the study due to incomplete data on independent and outcome variables, as well as covariates, resulting in a refined cohort of 1,198 individuals eligible for subsequent analysis (**Supplementary** Figure 1).

### Construction of mid-age LE8 score

We defined “middle age” as the age range of 45 to 65 years across exams 5-9 [17–19], due to the measurement of most LE8 components starting from exam 5. Considering participants might have undergone multiple examinations in this period, the average LE8 scores were calculated for these participants from exams 5-9.

The LE8 total score was derived based on the AHA guidelines [11], with modifications made to fit the FHS cohorts [20]. Briefly, individuals who quit smoking less than two years ago were given a score of 25 for smoking. The Dietary Approaches to Stop Hypertension (DASH) [21] diet score was calculated based on components including the intake of vegetables, fruits, nuts, legumes, and whole grains; as well as low-fat dairy, and the intake of red and processed meat, sugar-sweetened beverages, and sodium [22]. The physical activity score was computed by considering the duration and strength of activities including sleep, sedentary, and different strengths of activities (i.e. light, moderate, and vigorous) [23]. The other scores such as sleep quality, blood sugar levels, BMI, blood lipid, blood pressure were derived based on the standard of AHA [11]. We calculated the mid-age LE8 total score by averaging scores across all eight LE8 components. Additionally, two sub-scores, LE8 health behavior score and LE8 health factor score, were also obtained[11]. Furthermore, we classified the mid-age LE8 total score into three CVH categories: Poor-CVH (LE8 total score below 50), Intermediate-CVH (LE8 total score between 50 and 70), and Ideal-CVH (LE8 total score above 70) [11].

### Construction of dCDT scores

The details of the dCDT can be found in previous studies [12, 13, 24–26]. This study used the dCDT total score, and eight domain-specific scores including COM/COP Drawing Efficiency scores, COM/COP Simple Motor scores, COM/COP Information Processing scores, and COM/COP Spatial Reasoning scores.

### Diagnostic criteria of Alzheimer’s disease

The cognitive status of FHS participants has been consistently monitored using a variety of methods, including periodic assessments with the NP test battery [6, 27, 28]. To ensure accurate diagnoses, a dedicated team comprising neurologists and neuropsychologists reviewed all available information. This team reaches a consensus on whether participants meet the Diagnostic and Statistical Manual of Mental Disorders, 4th edition (DSM-IV) criteria for dementia and assigns specific subtype diagnoses as needed [29]. The diagnostic criteria for AD adhere to the National Institute of Neurological and Communicative Disorders and Stroke and the AD and Related Disorders Association [30].

### Covariates

Covariates included age at dCDT examination, delta age, sex, education levels, apolipoprotein-E (*APOE*) genotype. The delta age was calculated to show the difference in years between participants’ age at dCDT and their age at the most recent examination that established their mid-life period. Participants with *APOE* ε2/ε4 allele were excluded. The remaining participants were stratified into two groups based on the presence (ε3/ε4 and ε4/ε4) or absence (ε2/ε3, ε2/ε2 and ε3/ε3) of the ε4 allele.

### Statistical analysis

LE8 scores and dCDT scores were converted to z-scores. Linear regression models were used to examine the associations of dCDT total score with LE8 total score and CVH categories. Furthermore, the pairwise associations between LE8 sub-scores and dCDT sub-scores were analyzed by linear regression models (**Figure 1**). All models were adjusted for age at dCDT, sex, education level, and delta age. The multiple comparisons were adjusted by the false discovery rate (FDR) correction [31].

**Figure 1.**
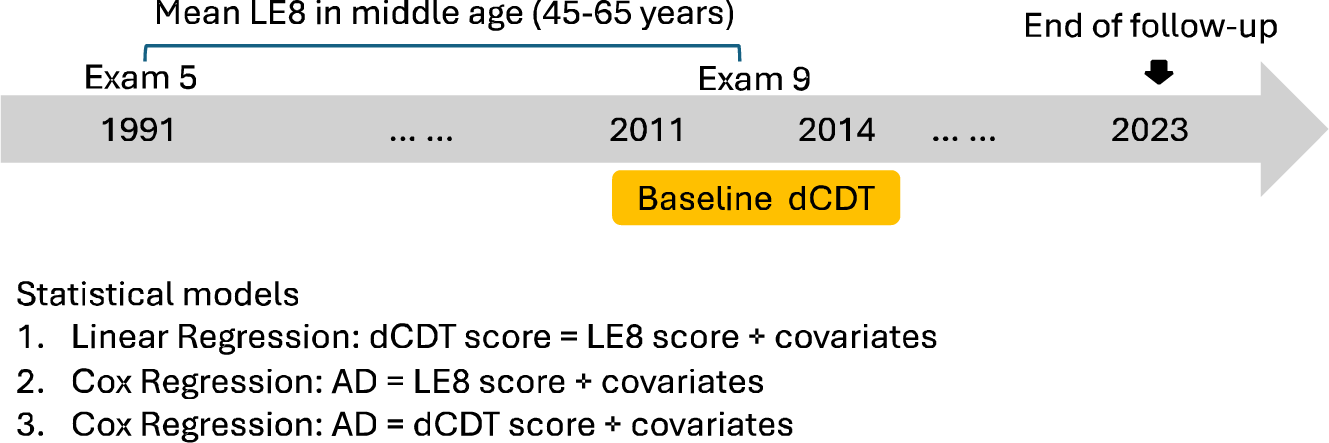
Study design. Middle age was defined as the age range from 45 to 65 years. Approximately 72% of participants in their middle age were assessed between Exam 5 and Exam 8 (from 1991 to 2008). We calculated the mean LE8 scores from all exams attended by a participant during their middle age. The statistical analyses included: 1. Association analyses of the baseline dCDT with the mean mid-age LE8 scores, adjusting for age at baseline dCDT, sex, and education, and the age difference between dCDT and the last LE8 measurement. 2. Association analyses of the mean mid-age LE8 scores with incident AD during a median follow up of 17.5 years, adjusted for age at the last LE8 measurement, sex, and education. 3. Association analyses of the baseline dCDT with incident AD during a median follow up of 8.9 years, adjusted for age at baseline dCDT, sex, and education.

Sex-specific analysis was performed to assess the variance in the effect size of the association between mid-age CVH and dCDT total score. Stratified analysis was further applied in *APOE* ε4 allele carriers and non-carriers. To determine whether sex and the presence of *APOE* ε4 allele modify the association between mid-age CVH and dCDT performance, we conducted additional analyses that include interaction terms (sex*CVH categories) and (*APOE* ε4 status*CVH categories) in the regression models, respectively.

Cox Proportional-Hazard model was used to examine the association between mid-age LE8 total score and incident AD. Adjustments were made for age at the time of the last FHS examination during middle age, sex, and education level (**Figure 1**). The follow-up period was determined from the end of middle age to the earliest occurrence of AD onset, death, or up to July 12, 2023. Additionally, we applied the Cox model using the categorical variable of mid-age CVH as the main predictor, while adjusting for the covariates.

The association between dCDT performance and incident AD was examined by Cox model with dCDT total score as primary predictor and AD incidence as the dependent outcome (**Figure 1**). The covariates included age at dCDT, sex, and education level. Follow-up time spanned from the dCDT assessment to the earliest occurrence of AD onset, death, or July 12, 2023. In the secondary analysis, we conducted Cox model with the eight dCDT subdomain scores as main predictors and AD onset as outcome, adjusting for the same set of covariates. We tested the proportional hazard assumption in all Cox models using the Schoenfeld residuals test.

We evaluated the predictive ability of the mid-age LE8 total score, dCDT total score, and various covariates for AD risk by constructing five logistic regression models and comparing their area under the receiver operating characteristic (ROC) curve (AUC). The baseline model included covariates with age, sex, and education level. Three subsequent models incorporated the following additional variables one at a time: mid-age LE8 total score, dCDT total score, and *APOE* ε4 status. The final comprehensive model encompassed age, sex, education level, mid-age LE8 total score, dCDT total score, and *APOE* ε4 status. All analysis were performed using RStudio version 4.2.1.

## RESULTS

### Participant characteristics

This study included 1,198 participants (mean age at dCDT measurement 73±7, 56.8% women, 47.5% college and higher). Forty-five participants were diagnosed with AD during the median of 17.5 years follow up. The detailed clinical characteristics were shown in **Table 1**.

**Table 1.**
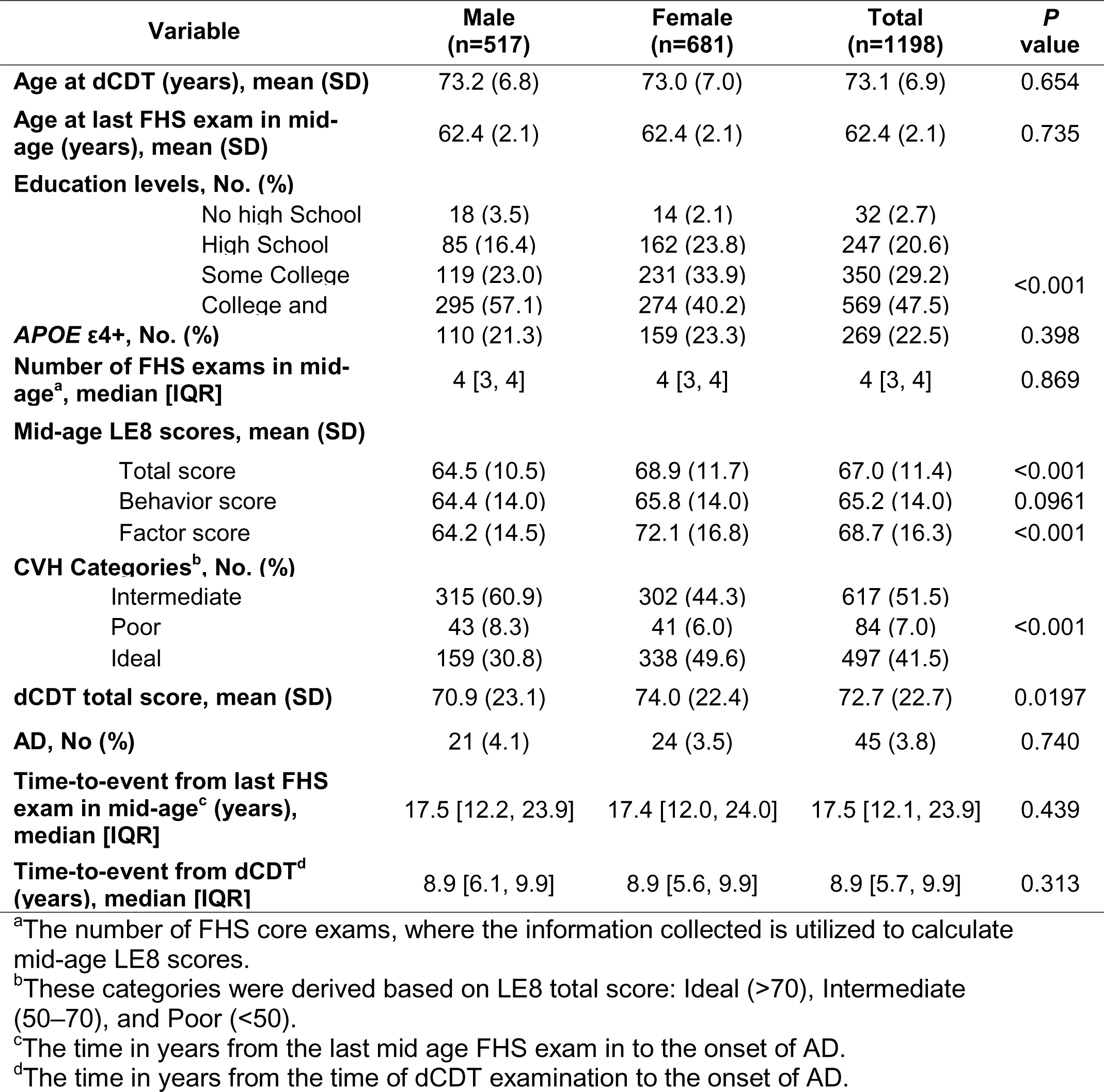
Characteristics of the 1198 participants in the FHS Offspring cohort

### Association of demographic variables with mid-age LE8 scores and dCDT scores

In the univariate analysis, with each additional year of age, participants’ dCDT total scores decreased by 0.07 SD (95% CI: -0.08, -0.06; *P*<0.001), indicating a decline in cognitive performance with age (**Supplementary Table 1**). Women demonstrated superior cognitive performance than men, scoring on average 0.19 SD (95% CI: 0.03, 0.36) higher on the dCDT total scores (*P*=0.02). Additionally, a clear educational gradient was observed in dCDT performance. No significant association was observed between education level and mid-age LE8 total score in the study participants (**Supplementary Table 2**).

### Association between mid-age LE8 scores and dCDT scores

An increase of one SD in the mid-life LE8 total score was positively associated with a 0.16 SD increase in the dCDT total score (95% CI: 0.06, 0.24; *P*<0.001) (**Figure 2)**. When using participants with intermediate CVH as a reference group, those within the ideal CVH showed a statistically significant increase of 0.18 SD in their dCDT total scores (95% CI: 0.01, 0.34; *P*=0.036). In contrast, those categorized with poor CVH had an average decrease of 0.36 SD in their dCDT total scores (95% CI: -0.67, -0.05; *P*=0.024) (**Table 2**).

**Figure 2.**
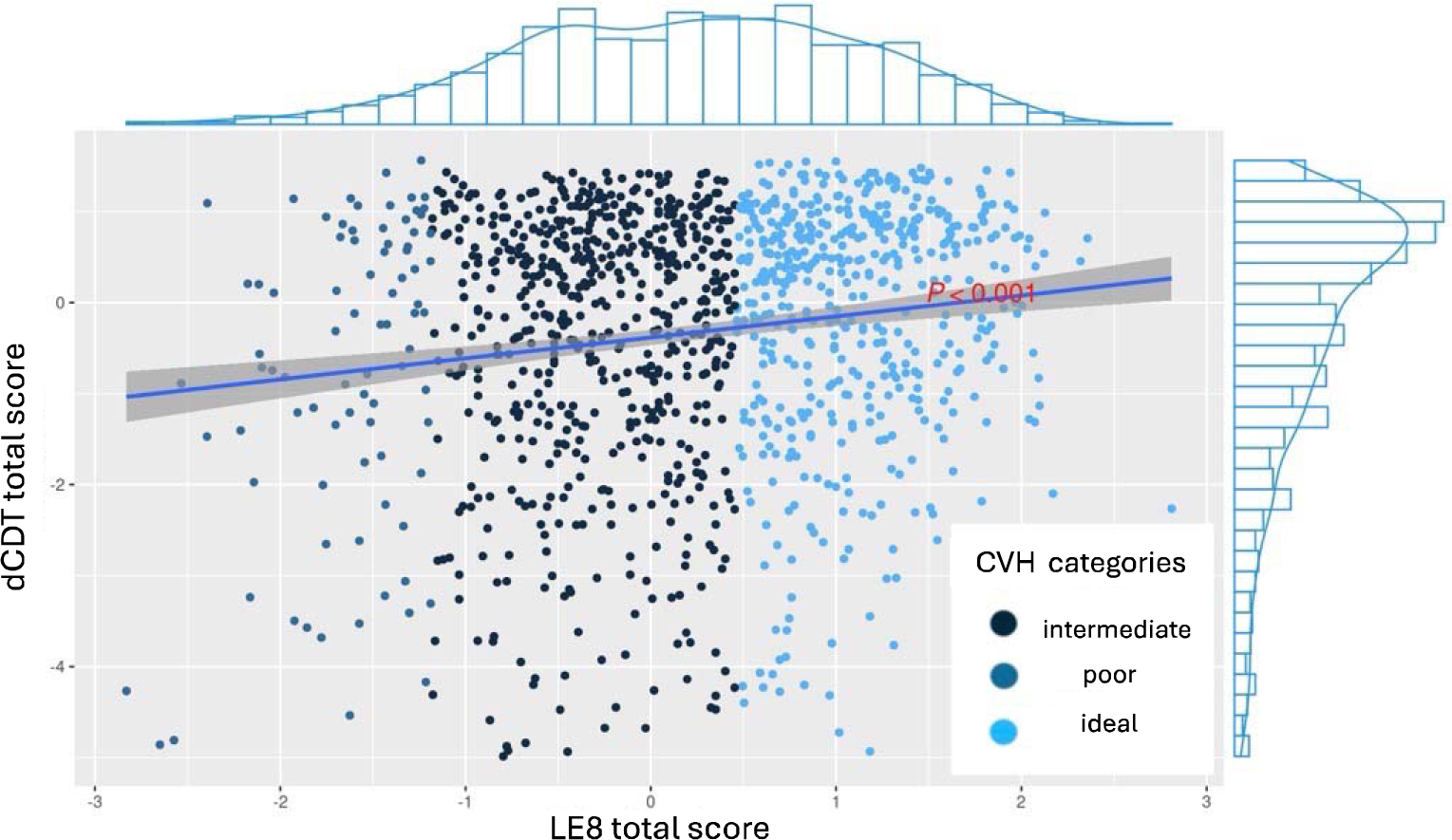
Association of CVH with dCDT total score and incident AD (n=1,198). A scatter plot with standardized mid-age LE8 total scores on the x-axis and standardized dCDT total scores on the y-axis, representing individual data points that are color-coded according to three levels of CVH: poor (light blue), intermediate (blue), and ideal (dark blue). A trend line is shown, indicating a positive correlation between LE8 total scores and dCDT total scores with statistically significant (*P*<0.0001). The distribution of LE8 total scores and dCDT total cores is illustrated by histograms.

**Figure 3.**
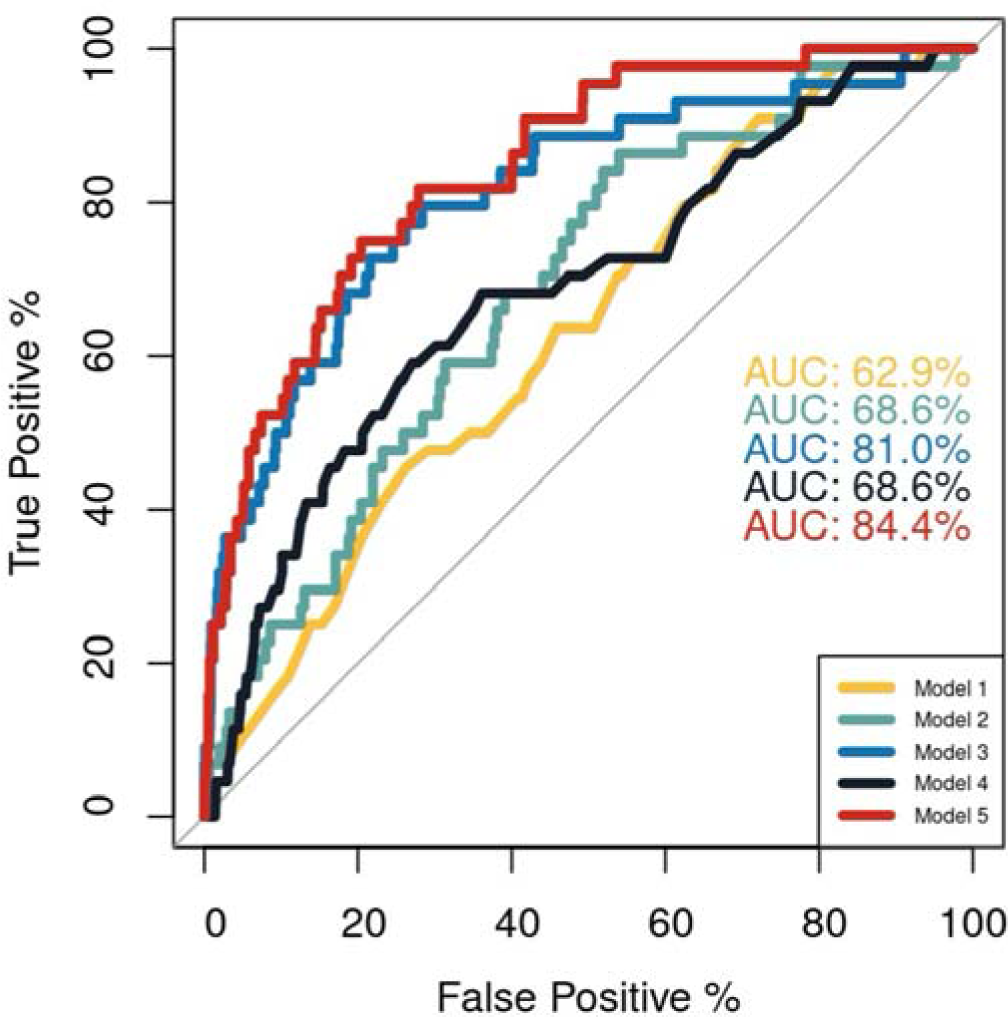
ROC curve plot that compares the AD prediction performance of five different models. Model 1: AD ∼ Age + Sex + Education; Model 2: AD ∼ LE8 total score + Age + Sex + Education; Model 3: AD ∼ dCDT total score + Age + Sex + Education; Model 4: AD ∼ *APOE* + Age + Sex + Education; Model 5: AD ∼ LE8 total score + dCDT total score + *APOE* +Age + Sex + Education

**Table 2.** Association of dCDT total score with LE8 total score and CVH categories

| **Main Predictor** | **Beta** | **95% CI** | **SE** | **P value** |
| --- | --- | --- | --- | --- |
| LE8 total score | 0.16 | (0.08, 0.24) | 0.04 | **<0.001** |
| CVH categories |  |  |  |  |
| Intermediate | Ref |  | - | - |
| Poor | -0.36 | (-0.67, -0.05) | 0.16 | **0.024** |
| Ideal | 0.18 | (0.01, 0.34) | 0.08 | **0.036** |

The association was examined by linear regression model adjusting for age at dCDT examination, delta age, sex, and education levels.

Higher LE8 total score were associated with enhanced dCDT performance (**Supplementary** Figure 2). The LE8 total score was significantly associated with most dCDT sub-scores (FDR-adjusted *P* ≤ 0.03), except for the scores of drawing efficiency and information processing under the command task. Compared to the LE8 health behavior score, the LE8 health factor score showed significant association with more dCDT sub-scores (4 versus 6) (**Supplementary** Figure 2).

### Effect modification by sex

An increase of one SD in the mid-life LE8 total score was significantly associated with a 0.20-SD increase in the dCDT total score among women (95% CI: 0.10, 0.30; *P*<0.001) (**Supplementary Table 3**). For men, this association did not achieve statistical significance (*P*=0.18). There was no significant interaction effect between mid-life LE8 total score and sex (*P*=0.195) (**Supplementary Table 4**). Women with poor CVH exhibited dCDT total scores that were, on average, 0.79 SD lower than those with intermediate CVH (95% CI: -1.22, -0.35; *P*<0.001) (**Supplementary Table 3**). This association was not observed in men. And the sex significantly influences the association between mid-life CVH categories and dCDT performance (**Supplementary** Figure 3), with a *P* value of 0.016 for interaction term.

### Effect modification by *APOE* **ε**4 status

No significant interaction effect of *APOE* ε4 status with mid-age LE8 scores (*P*=0.053) and CVH categories was observed (*P*=0.249) (**Supplementary Table 4**). There were variations in the strength of the association of dCDT scores with LE8 total score and the CVH categories when comparing *APOE* ε4 carriers to non-carriers (**Supplementary** Figure 4). Specifically, among *APOE* ε4 non-carriers, an increase in mid-life LE8 total score was associated with a 0.12 SD increase in dCDT total score (95% CI: 0.03, 0.21; *P*=0.009). In contrast, *APOE* ε4 carriers experienced a more pronounced improvement of effect size with a beta of 0.29 (95% CI: 0.11, 0.47; *P*=0.002) (**Supplementary Table 5**).

### Association of mid-age LE8 scores with incident AD

An increase of one SD in the mid-life LE8 total score was associated with a 45% reduction in the risk of incident AD (95% CI: 0.49, 0.87; *P*=0.003) (**Table 3**). Compared with participants with intermediate CVH, those with the ideal CVH appeared a 53% decreased AD risk (95% CI: 0.23, 0.98; *P*=0.045). Conversely, participants with poor CVH in mid-life were observed to have a 43% increased risk of incident AD (**Supplementary** Figure 5). For each SD increase in the mid-life LE8 behavior score, there was a 46% reduction in the risk of incident AD (95% CI: 0.49, 0.83; *P*<0.001) (**Supplementary Table 6**). The proportional hazards assumption was consistently satisfied in all Cox models.

**Table 3.** Association of incident AD with LE8 total score, CVH categories, and dCDT total score

| **Main Predictor** | **HR** | **95% CI** | **P value** |
| --- | --- | --- | --- |
| LE8 total score<sup>a</sup> | 0.65 | (0.49, 0.87) | **0.003** |
| CVH categories<sup>b</sup> |  |  |  |
| Intermediate | Ref | - | - |
| Poor | 1.43 | (0.59, 3.47) | 0.433 |
| Ideal | 0.47 | (0.23, 0.98) | **0.045** |
| dCDT total score<sup>c</sup> | 0.51 | (0.42, 0.61) | **<0.001** |

<sup>a</sup>The association of AD with mid-age LE8 total score (z-score) was examined by adjusting for age at last included FHS exam, sex, and education levels.
<sup>b</sup>The association of AD and CVH categories (intermediate was used as the reference group) was examined by adjusting for age at last included FHS exam, sex, and education levels.
<sup>c</sup>The association analyses of AD and dCDT total score (z-score) was examined by adjusting for age at dCDT, sex, and education levels.

### Association of dCDT scores with incident AD

Each SD increase in the dCDT total score was associated with a 49% lower risk of incident AD (95% CI: 0.42, 0.61; *P*<0.001) (**Table 3**). The significant associations between incident AD and domain-specific performances in all four domains under both command and copy tasks were observed (FDR-adjusted *P*<0.001) (**Supplementary Table 7**). Higher levels of domain-specific cognitive scores were associated with lower hazards of developing AD during the follow-up period, with HRs ranging from 0.51 to 0.67 (FDR-adjusted *P*<0.001).

### ROC Curve Analysis

Figure 4 illustrates the progression of predictive performance for incident AD starting from a base model that integrated age, sex, and education level, resulting in an AUC of 0.63. By incorporating the dCDT total score into this base model, the AUC increased to 0.81. Subsequent addition of both the LE8 total score and the *APOE* ε4 allele status to the base model each yielded AUCs of 0.69. The most comprehensive model, which included the dCDT total score, LE8 total score, and *APOE* ε4 status with the demographic variables, reached the highest AUC of 0.84.

## DISCUSSION

Our study investigated the association of mid-age LE8 scores with dCDT performance and incident AD. The results indicated individuals with higher LE8 total score in their middle ages tend to exhibit better cognitive performance and a reduced risk of incident AD. In addition, higher mid-age LE8 health factors and lifestyle behavior scores positively correlate specific cognitive functions in older age. The ROC analysis illustrated the model containing LE8 total score, dCDT total score, and *APOE* ε4 status achieved the highest AUC of 0.84 in predicting incident AD.

AD, as a neurodegenerative condition, may progress many years before noticeable symptoms occur.[32] Previous studies have underscored the importance of detecting and addressing risk factors in midlife in delay and prevention of AD in later years [33, 34]. Our study builds on this foundation by emphasizing the CVH in midlife as a critical period for implementing intervention and prevention strategies before the disease progresses significantly. Previous studies have established various methods of measuring CVH in middle age and analyzed the associations between mid-age CVH and AD [35, 36]. Another study discovered that mid-age Framingham Stroke Risk Profile was significantly associated with memory decline [37]. More studies have used LS7 to quantify CVH, and found that higher LS7 protect cognition [8, 9, 38–40]. LE8 incorporating eight modifiable risk factors and uses a 100-point scale for assessment, providing a more comprehensive and nuanced perspective of CVH evaluation [11]. Our study revealed consistent finding with a superior LE8 score in middle life was associated with improved cognitive performance and a reduced AD risk.

Digital cognitive assessment tests are becoming widely used and hold the potential to replace traditional paper-based tests as a rapid and more sensitive tool for detecting early cognitive decline. Establishing the relationship between CVH and these new modalities of cognitive performance is essential. A prior study that compared the performance of dCDT with conventional NP test observed that the composite scores built based on dCDT were superior surrogates for their corresponding NP tests [41]. Specifically, lower standard errors and more significant *P* values were found in the associations between dCDT composite scores and mild cognitive impairments compared with associations with NP tests [41].

Genetic factors, particularly the presence of the *APOE* ε4 allele, are known to significantly and negatively influence the onset of AD [42, 43]. Our study found that among *APOE* ε4 carriers, a correlation exists between CVH condition in middle age and a more pronounced cognitive decline, aligning with previously reported findings [38]. *APOE* ε4 carriers with suboptimal CVH in their middle ages tend to have a greater increase in the risk of AD. Conversely, *APOE* ε4 carriers with ideal CVH in this period are likely to see a more significant reduction in AD risk. Given the significant impact genetic factors, particularly the *APOE* ε4 allele, on AD onset and the observed correlation between mid-life CVH and cognitive decline among *APOE* ε4 carriers, we advocate for the adoption of healthy lifestyle habits and the maintenance of optimal CVH from an early age for all individuals. This approach is particularly emphasized for those with the *APOE* ε4 allele, given their increased risk, but is beneficial universally to mitigate AD risk.

Our study had several strengths. The FHS Offspring Cohort, being a longitudinal cohort, serves as a reliable and extensive data source. It uniquely offers both longitudinal LE8 measures and dCDT data, a rarity in other studies. Our study has limitations in several aspects. First, the FHS Offspring cohort comprises those who are non-Hispanic White from a relatively affluent town, many of whom have received a college degree or above education [35]. This may lead to limitations in the representability of our findings to more diverse populations. Additionally, we have a relatively small number of incident AD cases during a median of 17.5 years of follow-up since the middle age. The small sample size in AD poses challenges in conducting robust subgroup analysis. Therefore, further studies with diverse populations and larger sample sizes are warranted.

In summary, these results underline the importance of mid-life cardiovascular health in predicting cognitive outcomes and the development of AD. Our results indicated that optimal mid-age CVH is associated with enhanced cognitive function and a reduced risk of incident AD, particularly in women and those with higher AD genetic risk.

## Data Availability

The data used in this study could be requested through an application to the Framingham Heart Study.

https://www.framinghamheartstudy.org/fhs-for-researchers/

## ACKNOWLEDGEMENTS

We express our gratitude to the participants of the Framingham Heart Study for their commitment, acknowledging that this research could not have been conducted without their involvement. Additionally, we extend our thanks to the FHS researchers for their sustained dedication to subject examinations over the years.

## FUNDING

This work was supported by National Heart, Lung, and Blood Institute contract (N01-HC-25195; HHSN268201500001I), and NIH grants from the National Institute on Aging (AG008122, AG062109, AG068753)

## AUTHOR CONTRIBUTIONS

Jian Yang (Conceptualization; Methodology; Data Curation; Formal analysis; Investigation; Writing - Original Draft; Writing - Review & Editing; Visualization); Huitong Ding (Investigation; Writing - Original Draft; Writing - Review & Editing); Yi Li (Data Curation; Writing - Review & Editing); Ting Fang Alvin Ang, Sherral Devine, Yulin Liu, Wendy Qiu, Rhoda Au, and Jiantao Ma (Resources; Writing - Review & Editing; Other); Chunyu Liu (Conceptualization; Methodology; Writing - Original Draft; Writing - Review & Editing; Visualization; Supervision; Project administration; Funding acquisition; Other).

## CONFLICT OF INTEREST

Dr. Au is a scientific advisor to Signant Health and NovoNordisk, and a consultant to the Davos Alzheimer’s Collaborative. The other authors state that this study was carried out without any commercial or financial affiliations that might be seen as a possible conflict of interest.

## ETHICS APPROVAL AND CONSENT TO PARTICIPATE

The study’s procedures and protocols received approval from the Institutional Review Board at Boston University Medical Campus. All participants provided written informed consent.

## Notes

### Author Declarations

The institutional review board at Boston University gave ethical approval for this work.

