## Supplemental Tables and Figures for "Association of mid-age Life’s Essential 8 score with digital cognitive performance and incident Alzheimer’s disease: the Framingham Heart Study"

**Supplementary Table 1.** Univariate association analyses between each covariate and dCDT total score.

| **Predictors** | **Beta** | **95% CI** | **SE** | ***P* value** |
| --- | --- | --- | --- | --- |
| Age at dCDT | -0.072 | (-0.084, -0.061) | 0.006 | **<0.001** |
| Sex  Male  Female | Ref  0.193 | -  (0.027, 0.359) | -  0.085 | -  **0.023** |
| Education levels  No high school  High school  Some college  College and higher | Ref  0.445  0.668  0.972 | -  (-0.084, 0.974)  (0.148, 1.189)  (0.460, 1.483) | -  0.270  0.265  0.261 | -  0.099  **0.012**  **<0.001** |

**Supplementary Table 2.** Univariate association analyses between each covariate and mid-age LE8 total score.

| **Predictors** | **Beta** | **95% CI** | **SE** | ***P* value** |
| --- | --- | --- | --- | --- |
| Age at FHS exam^a^ | 0.0002 | (-0.027, 0.027) | 0.014 | 0.986 |
| Sex  Male  Female | Ref  0.383 | -  (0.271, 0.494) | -  0.057 | -  **<0.001** |
| Education Levels  No High school  High school  Some college  College and higher | Ref  -0.005  0.002  0.247 | -  (-0.368, 0.359)  (-0.355, 0.359)  (-0.104, 0.599) | -  0.185  0.182  0.179 | -  0.980  0.991  0.168 |

^a^Age at the last FHS exam in their middle age.

**Supplementary Table 3.** Stratification analysis of CVH in middle age and dCDT performance among sex groups.

| **Main Predictor** |  | **Male (n = 517)** | | |  | **Female (n = 681)** | | |
| --- | --- | --- | --- | --- | --- | --- | --- | --- |
|  | **Beta** | **95% CI** | **SE** | ***P* value** | **Beta** | **95% CI** | **SE** | ***P* value** |
| LE8 total score | 0.09 | (-0.04, 0.22) | 0.07 | 0.181 | 0.20 | (0.10, 0.30) | 0.05 | **<0.001** |
| CVH categories |  |  |  |  |  |  |  |  |
| Intermediate | Ref | - | - | - | - | - | - | - |
| Poor | 0.11 | (-0.33, 0.56) | 0.23 | 0.623 | -0.79 | (-1.22, -0.35) | 0.22 | **<0.001** |
| Ideal | 0.17 | (-0.10, 0.44) | 0.24 | 0.209 | 0.16 | (-0.04, 0.37) | 0.11 | 0.122 |

Note: All association analyses were adjusted for age at dCDT, education, and delta age.

**Supplementary Table 4.** Interaction analyses of sex and *APOE* ε4 status on association between mid-age CVH and dCDT total score in FHS Offspring participants (n = 1,198).

| **Interaction terms** | ***P* value** |
| --- | --- |
| LE8 total core*Sex^1^ | 0.195 |
| CVH category*Sex^2^ | **0.016** |
| LE8 total core**APOE* ε4 status^3^ | 0.053 |
| CVH category* *APOE* ε4 status^4^ | 0.249 |

Note: All interaction analyses were adjusted for age at dCDT, sex, education, and delta age.

**Supplementary Table 5.** Stratification analysis of CVH in middle age and dCDT performance among *APOE* ε4 status groups.

| **Main Predictor** |  | ***APOE* ε4 carrier (n = 269)** | | |  | ***APOE* ε4 non-carrier (n = 906)** | | |
| --- | --- | --- | --- | --- | --- | --- | --- | --- |
|  | **Beta** | **95% CI** | **SE** | ***P* value** | **Beta** | **95% CI** | **SE** | ***P* value** |
| LE8 total score | 0.29 | (0.11, 0.47) | 0.09 | **0.002** | 0.12 | (0.03, 0.21) | 0.05 | **0.009** |
| CVH categories |  |  |  |  |  |  |  |  |
| Intermediate | Ref | - | - | - | - | - | - | - |
| Poor | -0.66 | (-1.35, 0.04) | 0.35 | 0.065 | -0.26 | (-0.61, 0.08) | 0.18 | 0.140 |
| Ideal | 0.3 | (-0.08, 0.68) | 0.19 | 0.13 | 0.14 | (-0.04, 0.33) | 0.09 | 0.137 |

Note: All association analyses were adjusted for age at dCDT, education, and delta age.

**Supplementary Table 6.** Secondary association analysis between LE8 performance and incident AD.

| **Main predictor^a^** | **HR** | **95% CI** | ***P* value** |
| --- | --- | --- | --- |
| LE8 behavior score | 0.64 | (0.49, 0.83) | **<0.001** |
| LE8 health factor score | 0.84 | (0.63, 1.13) | 0.25 |

^a^We performed association analysis between mid-age LE8 performance and onset AD with each of the two LE8 sub-scores as predictor and AD incidence as outcome, adjusting for age at last FHS exam in middle age, sex, and education.

**Supplementary Table 7.** Secondary association analysis between dCDT performance and incident AD.

| **Tasks** | **Predictors** | **HR** | **95% CI** | ***P* value** |
| --- | --- | --- | --- | --- |
| Command Task | Drawing efficiency score | 0.67 | (0.56, 0.81) | **<0.001** |
|  | Simple motor score | 0.63 | (0.51, 0.77) | **<0.001** |
|  | Information processing score | 0.57 | (0.48, 0.67) | **<0.001** |
|  | Spatial reasoning score | 0.59 | (0.49, 0.70) | **<0.001** |
| Copy Task | Drawing efficiency score | 0.65 | (0.56, 0.76) | **<0.001** |
|  | Simple motor score | 0.57 | (0.46, 0.76) | **<0.001** |
|  | Information processing score | 0.56 | (0.46, 0.67) | **<0.001** |
|  | Spatial reasoning score | 0.51 | (0.41, 0.65) | **<0.001** |

Note: We applied association analysis between dCDT performance and onset AD with each of the subdomain scores as predictor and incident AD as outcome, adjusting for age at dCDT, sex, and education. All *P* values have been adjusted for false discovery rate (FDR).

**Supplementary Figure 1.** Flowchart of sample selection.

^a^One participant may have missing values for multiple variables.

**Supplementary Figure 2.** Heatmap of the secondary analysis between mid-age LE8 total and sub scores and dCDT total and sub scores. We applied association analyses using linear regression model to explore the relationships between each of the three mid-age LE8 scores and the nine dCDT scores within 1,198 participants from the FHS Offspring cohort. These analyses were adjusted for age at dCDT, sex, education, and delta age. The color intensity represents the t-values from these associations, while the annotated numbers indicate the FDR-adjusted p-values for each specific association.

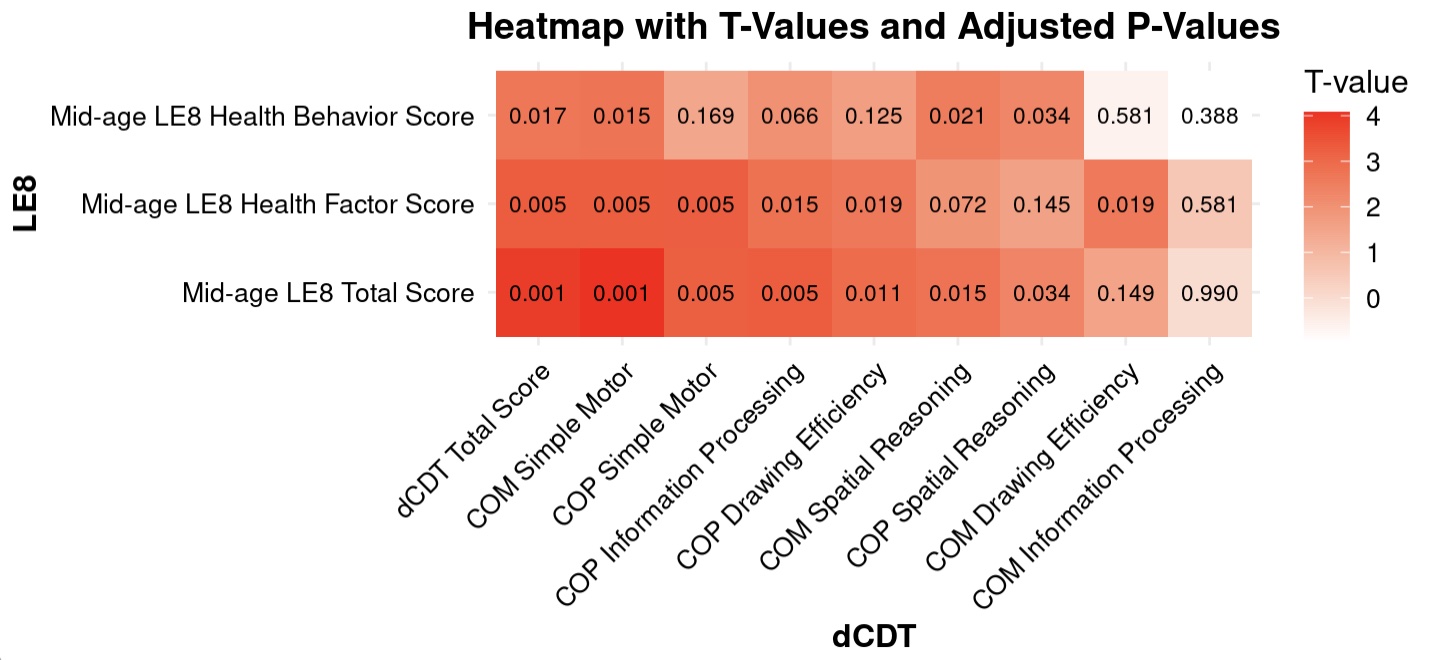

**Supplementary Figure 3**. Associative Interactions Between Mid-Age CVH and dCDT Performance by Sex. A scatter plot with mid-age LE8 total scores on the x-axis and dCDT total scores on the y-axis, representing individual data points that are color-coded with male colored in green and female colored in pink. Two crossed trend lines are shown, indicating a significant interaction of sex on the association between LE8 total scores and dCDT total scores.

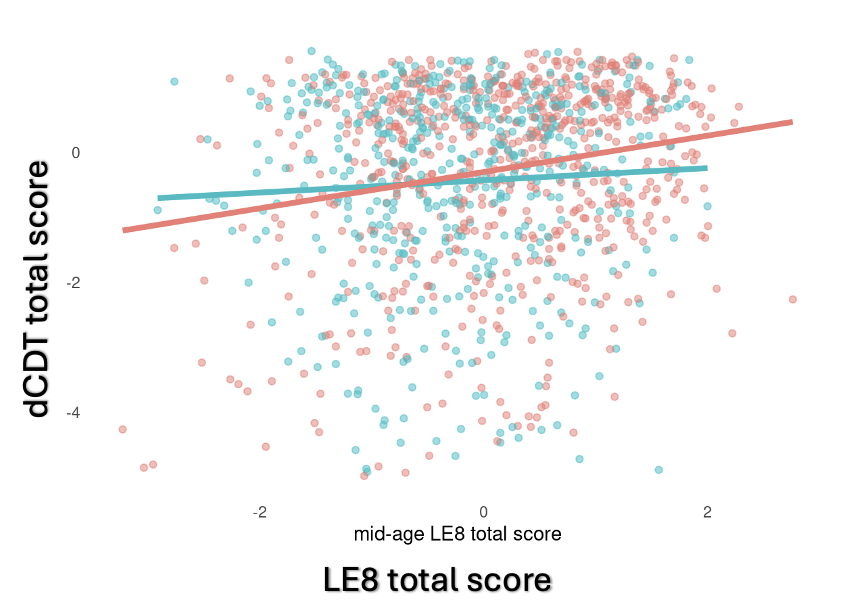

**Supplementary Figure 4**. Associative Interactions Between Mid-Age CVH and dCDT Performance by *APOE* ε4 Status. A scatter plot with mid-age LE8 total scores on the x-axis and dCDT total scores on the y-axis, representing individual data points that are color-coded with non-*APOE* ε4 allele carriers colored in red and *APOE* ε4 allele carriers colored in blue. Two crossed trend lines are shown, indicating a significant interaction of *APOE* ε4 allele status on the association between LE8 total scores and dCDT total scores.

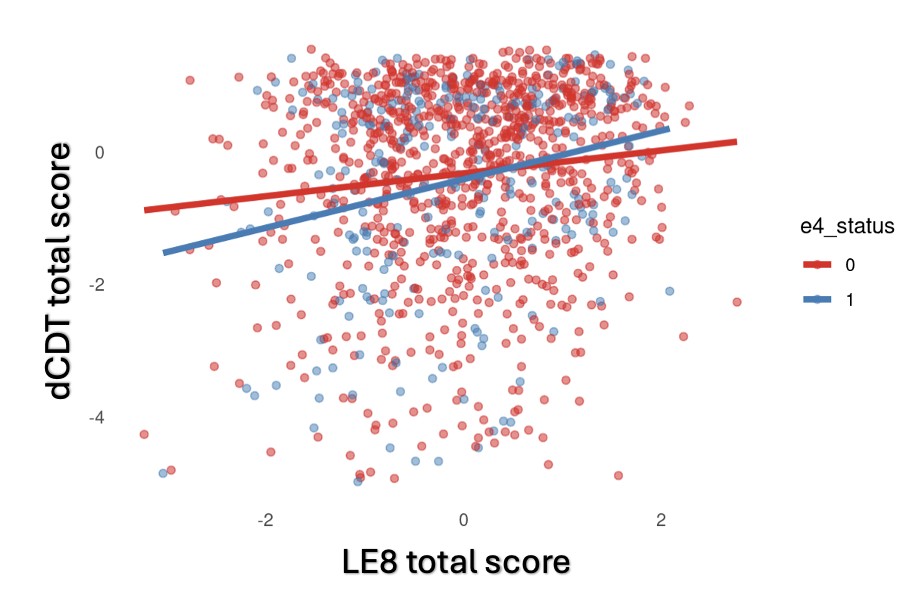

**Supplementary Figure 5**. Association of CVH with dCDT total score and incident AD (n=1,198). A forest plot illustrating the hazard ratios for different categories of CVH in middle age. On the x-axis, there are three mid-age CVH categories: ideal, intermediate, and poor. The y-axis represents the hazard ratio. Each CVH category has a corresponding point estimate of the hazard ratio with vertical lines representing the 95% confidence intervals.

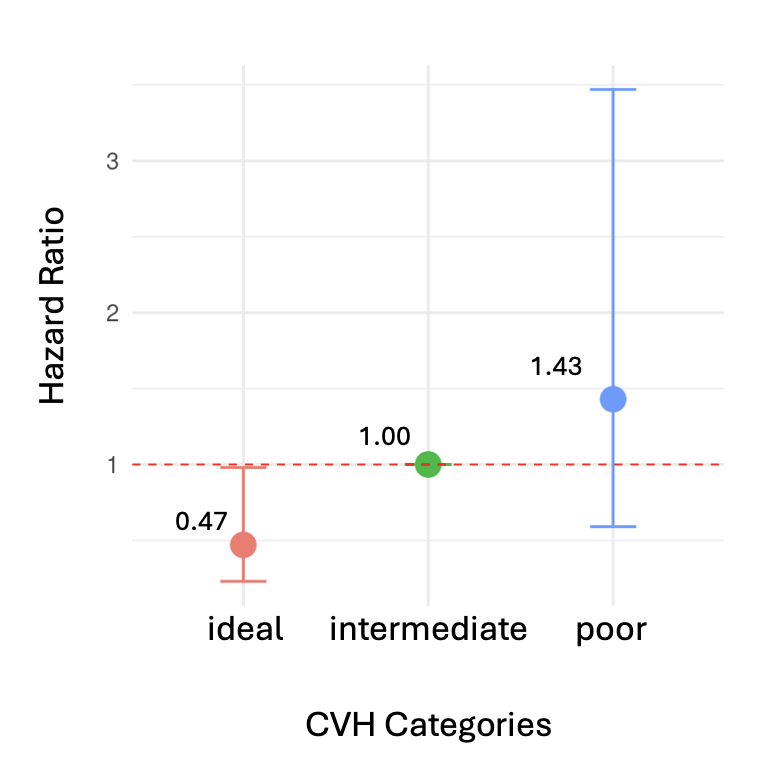
